# Effects of the ‘Reac Step’ training program on balance recovery and fall risk factors in older people: An assessor-blinded randomised controlled trial

**DOI:** 10.64898/2026.02.18.26346028

**Authors:** Shivam Sharma, Carly Chaplin, Cameron Hicks, Daniel Treacy, Melanie Farlie, Steven Phu, Natassia Smith, Hannah Gibson, Chrissie Ho, Eve Coleman, Tiffany Huang, Colleen G Canning, Natalie E Allen, Jaqueline T Close, Stephen R Lord, Yoshiro Okubo

**Author notes:** **Corresponding author** Yoshiro Okubo, PhD, Neuroscience Research Australia, 139 Barker Street, Randwick, New South Wales, Australia 2031. **Contribution Statement** Y.O., C.Ho., T.H. and S.R.L. contributed to the conception and design of the study. Y.O., C.C., C.Hi., C.Ho., N.S., H.G., E.C., S.P. and T.H. administrated the study. C.H. conducted the statistical analyses. The manuscript was drafted by S.S. and Y.O. and reviewed and edited by C.G.C., N.E.A., C.C., M.F, C.Hi., C.Ho., N.S., H.G., E.C., S.P., T.H., D.T, J.T.C. and S.R.L.

## Abstract

**Background:** Reactive balance training using repeated perturbations may reduce falls, however, training methods are not easily replicated or translatable to clinical settings. This study aimed to examine the effects of a novel reactive balance training program on balance recovery from laboratory induced trips and slips and fall risk factors in older people using simple and low-cost equipment.

**Methods:** We conducted a randomised controlled trial involving 88 older people. An intervention group (n = 43) received the ReacStep program which involved tether-release reactive stepping and intentional slips once a week for 6 weeks. Both the intervention and control (n = 45) groups received home-based strength training for 8 weeks. Blinded staff assessed reactive balance (laboratory induced falls), physical functions at baseline (week 1) and post intervention (week 8). Weekly SMS surveys ascertained falls in daily life over 12 months.

**Results:** Both groups were comparable in demographics, with a mean age of 72 years (SD = 5.6). Adherence to ReacStep sessions was high (90%). There were no significant differences between groups in laboratory-assessed reactive balance falls at post-test or daily-life falls over one year (*P* =.19). However, at post-test, the intervention group demonstrated significant improvements in usual gait speed, maximum step length, and choice stepping reaction time compared to controls (*<u>P</u>* <.05).

**Conclusions:** The ReacStep program demonstrated excellent adherence, was well tolerated, and improved gait parameters required for balance recovery following postural perturbations in older people. Nevertheless, it appears this program is not sufficient to improve reactive balance against unexpected trips and slips.

**Key points:**

- The ReacStep program is acceptable, demonstrates excellent adherence and improves gait measures in older people, potentially reducing fall risk.
- The generalisability against unexpected trips, and slips, and falls in daily life may be limited.
- Future research should explore more ecological perturbations while maintaining its accessibility and acceptability.

## INTRODUCTION

Traditional fall-prevention exercise programs require a high dose, around three hours per week, of challenging balance exercises to reduce falls by up to 39% in older adults (1). However, these programs may not equip older adults to recover from unexpected perturbations such as slips and trips during daily activities.

Reactive balance training (RBT), also known as perturbation-based balance training, utilises repeated postural perturbations to specifically improve reactive balance (2). During RBT, participants must integrate tactile, proprioceptive, visual, somatosensory, and vestibular inputs to detect postural threats from sudden external perturbations and restore stability by executing rapid, compensatory steps (1). Systematic reviews of randomised controlled trials (RCTs) have shown that RBT improves balance recovery skills and reduces falls in daily life by approximately 50% (2, 3). However, RBT requires sophisticated perturbation equipment such as perturbation treadmills (e.g. belt accelerations) (4), walkways with moveable plates and/or obstacle pop-up systems (5) which are not easily accessible. There are also safety concerns in exposing older adults to sudden and unpredictable perturbations (6). Higher rates of adverse events are reported in RBT (29%) compared to control exercises without perturbations (20%) (7). Manual perturbations applied by a therapist (e.g. pulls) (8) have advantages for clinical feasibility, but their effectiveness is unclear because their task-specificity to daily-life hazards may be limited.

An intermediate program that incorporates key aspects of the sophisticated interventions without requiring expensive laboratory set-ups may offer a practical, efficacious solution. This could include tether-release reactive step training - a simple, low-cost method that enables progressive intensity (9). Additionally, a recent pilot RCT in young adults found that high-dose volitional slip training using a visible hazard was as effective as low-dose unpredictable slip training with an undetectable hazard for improving slip recovery (10). Such volitional training provides high kinematic task-specificity (e.g., shaping correct step recovery responses) but lacks unpredictability, which may benefit individuals with balance impairments who fear falling, as unpredictability is a major source of anxiety during RBT (5). Finally because muscle weakness can limit balance recovery (5, 11), incorporating strength training targeting muscles used in reactive stepping may further enhance RBT effects (12).

Building on the above findings, we developed ReacStep - a task-specific program combining tether-release reactive step training, volitional slip training, and functional strength training, prioritizing safety, acceptability, and ecological validity (9). This study aimed to evaluate the effects of this novel, low-cost program on balance recovery from laboratory-induced trips and slips, as well as fall risk factors in older adults. We hypothesized that a 6-week ReacStep program would improve the ability to recover from unexpected trips and slips and improve gait, reaction time, and stepping performance.

## METHODS

### Design, ethics and registration

This study was an assessor-blind, two-arm, parallel-superiority RCT with a 1:1 allocation ratio. It was conducted from August 2022 till February 2024. The trial protocol was approved by the Institute Human Research Ethics Committee (HC210350) and prospectively registered on clinical trial registry.

### Study participants

Participants were recruited via the research volunteer (HC14320), research centre participant database, advertisements to seniors groups, clinics, hospitals, community groups, websites, institutions, and newspapers and social media. Eligibility criteria were as follows: aged 65 years and over, living independently in the community, able to walk 50 metres without mobility aid or resting. The exclusion criteria comprised diagnosed neurological diseases (e.g. Parkinson’s disease, multiple sclerosis, and/or dementia), existing conditions that prevent exercise (e.g. severe pain, heel ulcers, exercise intolerance), medical advice against exercise, history of lower limb, pelvis or vertebral fracture (s) or lower limb joint replacement(s) in the past 6 months. All participants provided informed consent prior to study participation.

### Sample size

A priori power analysis using G*Power 3.1.9.2 showed that 90 participants would provide 80% power to detect a medium-sized between-group difference (d =.6) in physical function parameters (2) with an independent t-test, assuming an alpha level of 0.05 and a 1:1 allocation ratio.

### Randomisation and blinding

The participants were randomly assigned to the intervention or control group using randomisation software (Blinders) with random block sizes. To ensure concealment and blinding, randomisation was conducted following baseline assessments (by C.C and Y.O), and post assessments were conducted by research assistants blinded to group allocation. To mitigate the placebo effect, both groups were provided with home-based strength training. (Figure 1)

**Figure 1.**
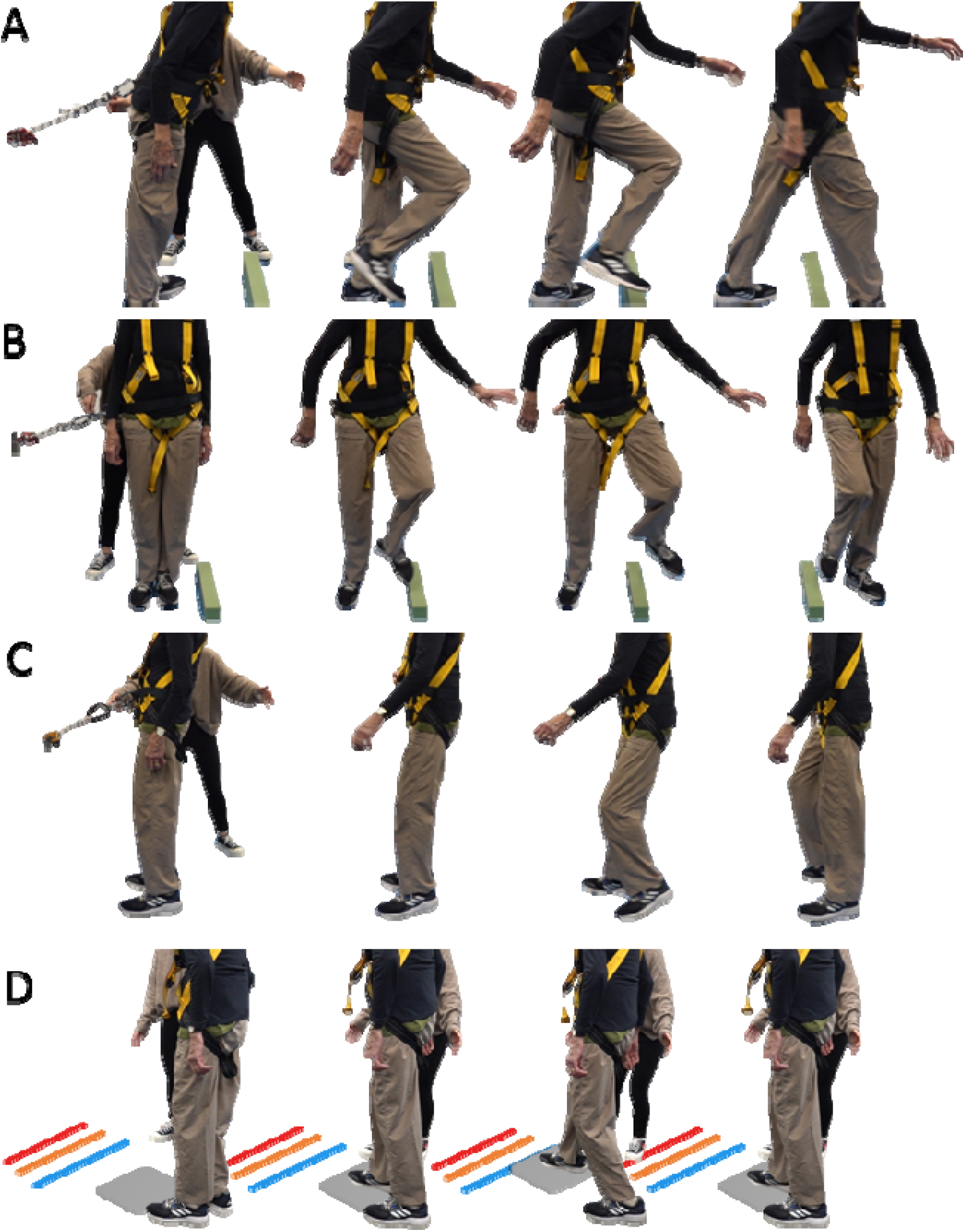
(A) Reactive step training in the forward direction. (B) sideways (C) backward direction (D) volitional slip training.

### Interventions

Participants in the intervention group completed a 45-minute ReacStep training session weekly for 6 weeks (from week 2 to week 7), supervised by an Exercise Physiologist (EP). Each session included a warm-up (∼5 minutes), reactive step training (∼30 minutes), and volitional slip training (∼10 minutes).

### Reactive Step Training

#### Tether-release training

For trips, tether-release reactive step training (Figure 1A) involved 5-20 trials per foot (left and right) in forward, lateral, and backward directions (40-80 trials total). Participants leaned against a rope anchored to a wall, initially loaded to ∼5% of body weight. The rope was released at random intervals (1-5 s), prompting a reactive step over a foam block (7 or 14 cm height) to regain balance within two steps (Figure 2A, B and C). Tether loading was progressively increased to a maximum of 20% of body weight as performance improved. A fall-arrest harness was used during training sessions.

**Figure 2.**
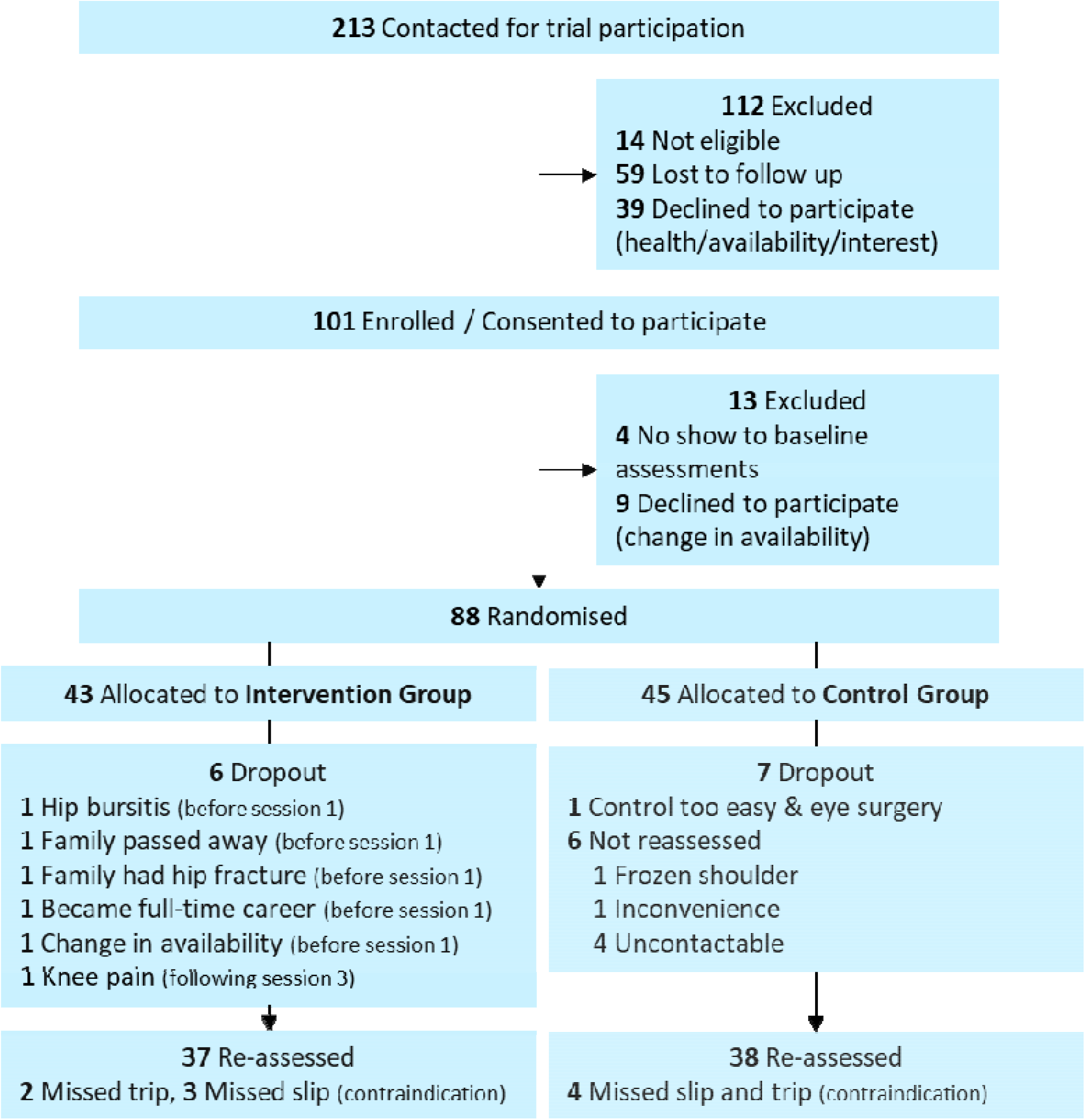
Trial flow chart

After the first trial in each direction, a cognitive dual task was introduced: participants memorised one word per trial while leaning and recalled the words after each release. By the tenth trial, they were required to recite all nine words in sequence.

#### Volitional Slip Training

Volitional slip training started with a standing position, participants stepped with one foot onto a hard plastic sheet (PVC, 50 × 50 cm) sprayed with silicone (Figure 1B) and pushed it forward to one of three target lines (20, 40, or 60 cm), then pulled it back while keeping the rear foot planted. These single-step slips were repeated 5 times per foot for each target (30 trials). Participants then took an additional step before stepping onto the sheet to increase momentum and slip speed, challenging more dynamic balance while slipping. These two-step slips were repeated five times per foot for each target (30 trials). Each session included approximately 60 volitional slips (Figure 2D).

### Home exercises

Both groups were provided with a set of resistance bands (66fit Resistance Band Exercise Loops with 4-5 levels of resistance) and completed a home-based lower limb strength program. Exercises included high knee marches and squats (targeting quadriceps, gluteals, and hamstrings), standing hip abductions (targeting hip abductors), and glute kickbacks (targeting gluteals and hamstrings) for 10 repetitions and 2 sets. In addition to a demonstration by an EP, a <u>video demonstration</u> was provided, and participants were instructed to perform the exercises twice weekly from week 1 to week 8.

### Control group

Control group received similar home-based exercises.

### Assessments

#### Timing and Blinding

Assessments were conducted by blinded assessors at baseline (week 1) and post-intervention (week 8).

#### Baseline Measures

Anthropometrics, age, gender, education, falls in the past 12 months, and subjective memory complaints were recorded.

#### Primary Outcome: Reactive Balance

The rate of falls following induced trip and slip in a laboratory setting was assessed using an 8-m Trip and Slip Walkway at post-assessment (5). A fall was defined as a harness-supported load exceeding 30% of body weight (13). Participants walked to match the bet of a metronome (set to 95% of their usual cadence) and stepped onto target tiles (set to 95% of their normal step length) to approximately 90% of normal gait speed. This measurement was performed with a blinded assessor. After 5–10 familiarisation walks, perturbations were introduced in a fixed but undisclosed order: a 70 cm slip (right foot) and a 14 cm obstacle trip (left foot), with at least one washout walk between trials.

#### Physical Function

Maximum step length (cm) was assessed with a step-in forward, backwards, left, and right directions with arms crossed at the chest over three trials (14). Maximal isometric knee extension strength (kg) was measured on the dominant leg using a dynamometer while seated across three trials, The best performance of the three recorded as the maximal strength measure (15). Postural sway path (mm/30 s) was recorded using a sway meter at waist level while standing on a foam block for 30 seconds (16). Choice Stepping Reaction Time (CSRT) (ms) was assessed using a visual display and a custom step mat (150 × 90 cm) with six stepping panels (2). Usual gait speed (m/s) was measured over 10 m across three trials on a 12-m course with 1-m acceleration/deceleration zones using a stopwatch. Dual-task gait speed was measured on the same course while counting backwards by threes.

#### Cognition, psychology and behaviours

Executive function was assessed by connecting numbers (A) and then alternating numbers and letters (B) with the Trail Making Test (TMT) (s) (17). Processing speed, scanning and attention were measured by the Symbol Digit Modalities Test (SDMT), requiring participants to match numbers to symbols using a reference key within 90 seconds (correct responses/90 s).(18). Short- and long-term verbal memory were evaluated using the Hopkins Verbal Learning Test-Revised (HVLT-R) (19). The Falls Behavioural Scale (FaB) was used to measure participant’s awareness and practice of behaviours associated with fall risk in daily life (20).

#### Daily falls and near-fall experiences

Falls, trips, and slips in daily life were prospectively recorded via weekly SMS surveys (REDCap/Twilio) and a falls diary for 52 weeks post-randomisation. A fall was defined as an unexpected event resulting in coming to rest on the ground, floor, or lower level (21). A trip was defined as catching the foot on an object causing imbalance (without falling), and a slip as the foot sliding on a surface causing imbalance (without falling) (22).

#### Acceptability and safety

Adherence to the ReacStep program and home-based training was monitored using a training log and weekly SMS surveys, respectively. Training progress was quantified by recording the maximum tether loading (% body weight) in successfully recovered trials across forward, lateral, and backward directions. Adverse events during ReacStep were monitored by the EPs. Exercise enjoyment was assessed using the Physical Activity Enjoyment Scale (PACES), adapted to reflect the exercises included in this study. Anxiety and perceive difficulty were measured via Likert scale.

#### Statistical Analysis

Analyses were conducted using an intention-to-treat approach. Post-intervention physical, cognitive, and enjoyment outcomes were compared using analysis of covariance (ANCOVA), adjusting for baseline values (except enjoyment, which had no baseline). Trip- or slip-induced falls in the laboratory were analysed using multiple logistic regression, with group allocation as the independent variable and age and gender as covariates. Daily life falls, trips, and slips were analysed using negative binomial regression with the same covariates. Changes in anxiety, perceived difficulty, and tether loading between the first and last ReacStep sessions were assessed using paired *t*-tests. All analyses were performed using IBM SPSS Statistics v29 *(IBM Corp*., *NY, USA)*, with statistical significance set at *P* <.05.

## RESULTS

### Participant Characteristics

Of the 101 individuals who expressed interest in the study, 88 completed baseline assessments and were subsequently randomised into the intervention (n = 43) and control (n = 45) groups. Seventy-five participants were re-assessed and included in the final analysis (intervention: n = 37; control: n = 38) (Figure 2). There were no significant differences in baseline characteristics between those who were re-assessed (n = 75) and those who were not (n = 13) (*P* >.05; Supplementary Table 1). Additionally, no significant differences were observed between the intervention and control groups in terms of age, gender, anthropometry, pain, fear of falling, or fall history (Table 1).

**Table 1.** Baseline participant characteristics (N=88)

| Variables | Intervention group<br>(N=43) | Control group<br>(N=45) |
| --- | --- | --- |
| Age (years) | 73.2 (6.1) | 71.6 (5.7) |
| Gender (female) | 65% (28) | 76% (34) |
| Height (cm) | 167.6 (10.1) | 165.6 (8.0) |
| Weight (kg) | 73.0 (14.6) | 71.0 (14.2) |
| BMI (kg/m<sup>2</sup>) | 26.0 (4.5) | 25.9 (4.9) |
| Osteoporosis (yes) | 16% (6) | 16% (7) |
| Back pain (yes) | 14% (5) | 26% (11) |
| Hip pain (yes) | 24% (9) | 12% (5) |
| Knee pain (yes) | 30% (11) | 26% (11) |
| Feet pain (yes) | 19% (7) | 12% (5) |
| Anxiety disorder (yes) | 11% (4) | 2% (1) |
| Walking stick (use) | 2% (1) | 2% (1) |
| Fear of falling (yes) | 81% (35) | 80% (35) |
| Falls in the past year (1+ falls) | 42% (18) | 45% (20) |

Values are means (standard deviations) or prevalence % (N). Fear of falling include mild, moderate and extreme. BMI: Body mass index

### Reactive balance (laboratory-induced trips and slips) and daily fall rate

Following a laboratory trip perturbation, 55.9% (n=19) of the intervention group and 60.6% (n=20) of the control group fell, with no statistical difference between groups (OR = 0.82; 95% CI, 0.31-2.18; *P* =.695). Following a laboratory slip perturbation, 57.6% (n=19) of the intervention group and 66.7% (n=22) of the control group fell, with no statistically significant difference (OR = 0.68; 95% CI, 0.25-1.85; *P* =.447). At 12 month follow-up, a total of 108 falls were reported. There was no between-group difference in the rate of falls in daily life (RR = 1.48; 95% CI, 0.81-2.69; *P* =.199). The intervention group reported more trip encounters(near-fall) than the control group (RR = 3.93; 95% CI, 1.73-8.03; *P* =.001), whereas no significant group difference was found for slip encounters (*P* =.123). (Table 2)

**Table 2:**
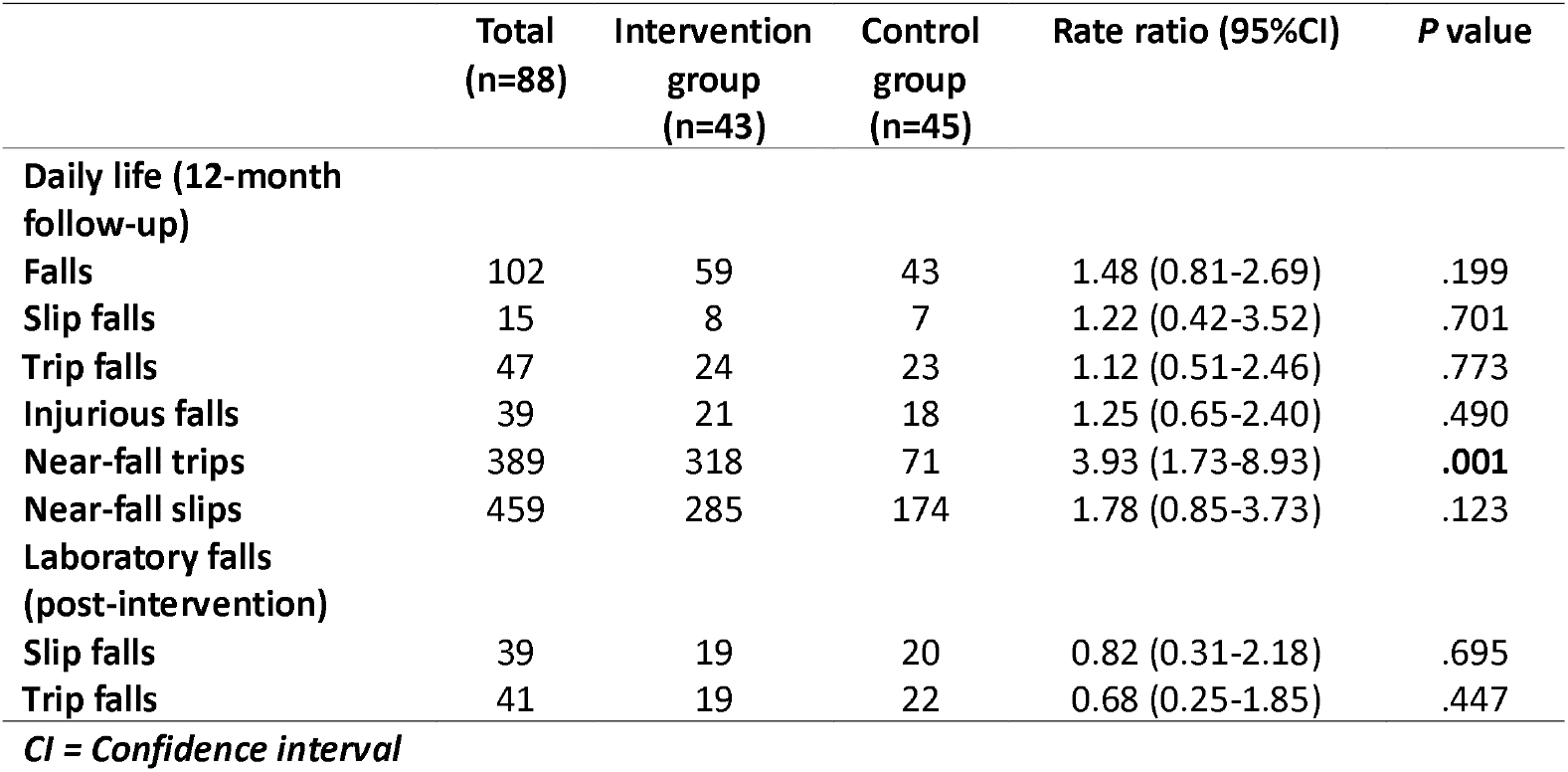
Falls results.

### Adherence and Adverse Events

Five participants from the intervention group withdrew prior to commencing training due to medical issues or personal circumstances (Figure 2). Among those who commenced training, 33 participants (90%) attended at least five of the six scheduled sessions. The most common reasons for non-attendance included COVID-19 infection (43%), travel (19%), and body pain (19%).

Home-exercise adherence was high, with 92% of participants in both groups completing at least 13 of the 16 prescribed sessions over the 8-week period. Three participants with pre-existing lower limb pain reported increased discomfort during the ReacStep intervention. One participant withdrew, while the remaining two completed the program with reduced training intensity and/or dosage. Three adverse events occurred during assessments: two participants sustained muscle strains (neck and lower back) following trip/slip perturbations, and one experienced a muscle strain during the knee extension strength assessment test.

### Physical and cognitive functions

There were no baseline between-group differences in any physical and cognitive measures (*P* >.05) (Table 3). Following the 6-week intervention and compared to the control group, the intervention group had higher usual gait speed (mean difference = 0.09 m/s; 95% CI, 0.02 - 0.16; *P* =.013, d =.3), longer maximum step length in the forward (mean difference = 9.18 cm; 95% CI, 3.68-14.67; *P* =.001, d =.5) and right (mean difference = 4.22 cm; 95% CI, 0.25-8.18; *P* =.037, d =.4) directions, shorter CSRT total time (mean difference = –72.80 ms; 95% CI, –137.4 to –8.15; *P* =.031, d = –.4) and lower FaB scores (mean difference = –0.10; 95% CI, –0.18 to –0.01; *P* =.023, d = –.1). No other variables showed significant between-group differences at the post-assessment.

**Table 3.**
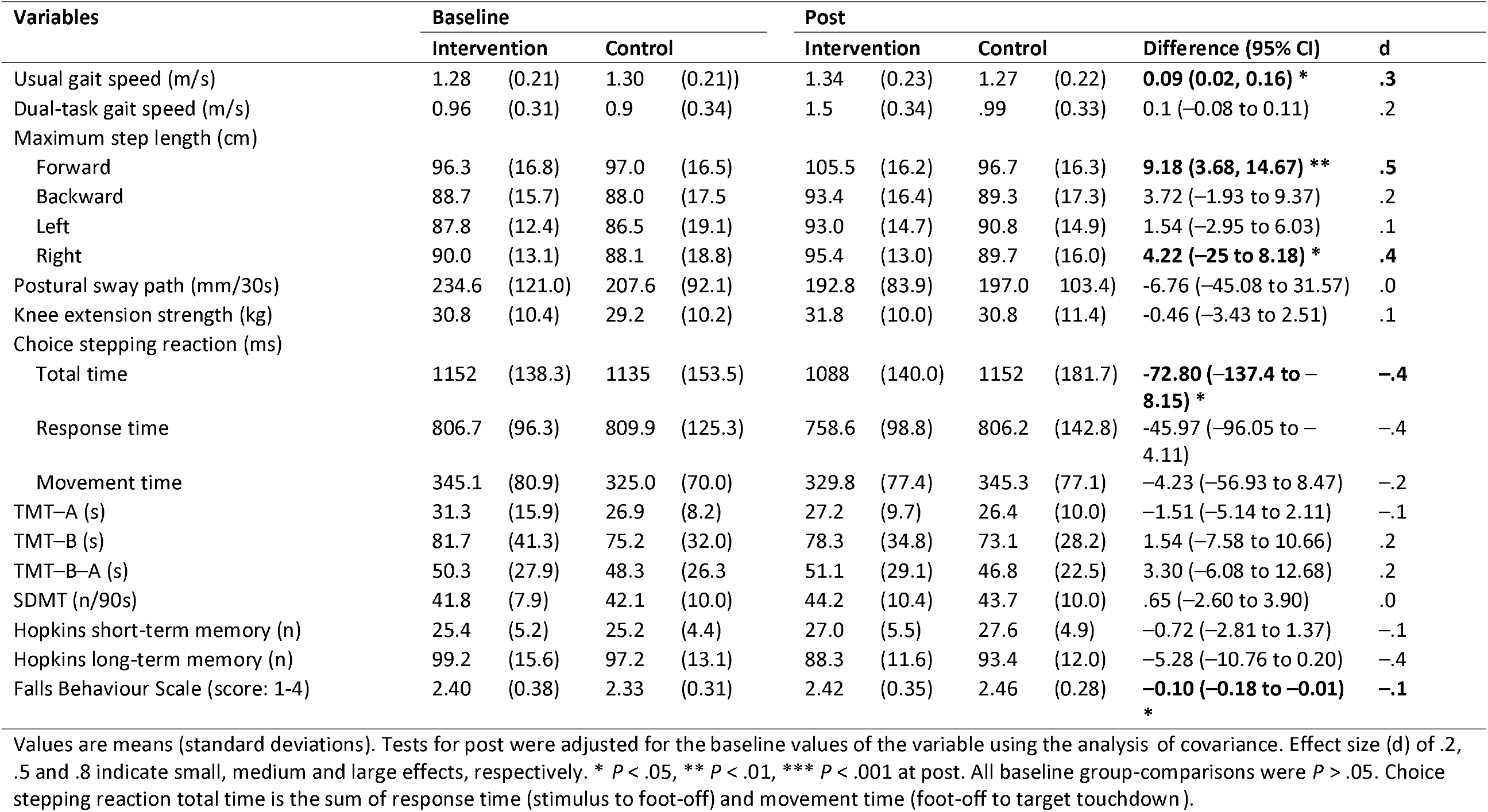
Physical and cognitive function measures at baseline (week 1) and post-intervention (week 8) (N=75)

### Training progression, anxiety, perceived difficulty and enjoyment

In the intervention group, maximum tether loading during reactive step training increased significantly across all directions between session 1 and session 6: forward (3.6% [12.7], 5.5% [17.3]), lateral (4.1% [9.6], 5.1% [14.0]) and backward (4.7% [9.5], 5.1% [14.6]) directions (*P* <.001). There was also a significant reduction in anxiety (95% CI, 0.49-0.86; *P* <.05, d =.46) and perceived difficulty (95% CI, 0.20-1.04; *P* <.01, d =.62) (Figure 3). Enjoyment of the exercise was significantly higher in the intervention group compared to the control group across all variables (*P* <.05) (Table 4)

**Table 4.** Enjoyment of the exercise programs for the intervention (ReacStep plus strength training) and control (strength training only) groups (N=72)

| Variables | Intervention group<br>(n=34) | Control group (n=38) | *P* | *d* |
| --- | --- | --- | --- | --- |
| Pleasurable | 5.8 (1.4) | 5.0 (1.3) | **.016** | .6 |
| Fun | 5.4 (1.4) | 4.4 (0.4) | **.002** | .8 |
| Pleasant | 5.4 (1.4) | 4.6 (1.5) | **.012** | .6 |
| Invigorating | 5.3 (1.3) | 4.6 (1.5) | **.023** | .5 |
| Gratifying | 5.6 (1.3) | 4.7 (1.4) | **.009** | .6 |
| Exhilarating | 4.4 (1.4) | 3.5 (1.6) | **.013** | .6 |
| Stimulating | 5.1 (1.4) | 4.2 (1.3) | **.008** | .6 |
| Refreshing | 4.5 (1.6) | 4.2 (1.4) | .343 | .2 |
| Total enjoyment | 41.5 (8.5) | 35.1 (7.8) | **.001** | .8 |

Values are means (standard deviations). Effect size (Cohen's *d*) of .2, .5 and .8 indicate small, medium and large effects, respectively. Higher scores indicate higher enjoyment ranging from 1-7 for each item and 8-56 to total.

**Figure 3.**
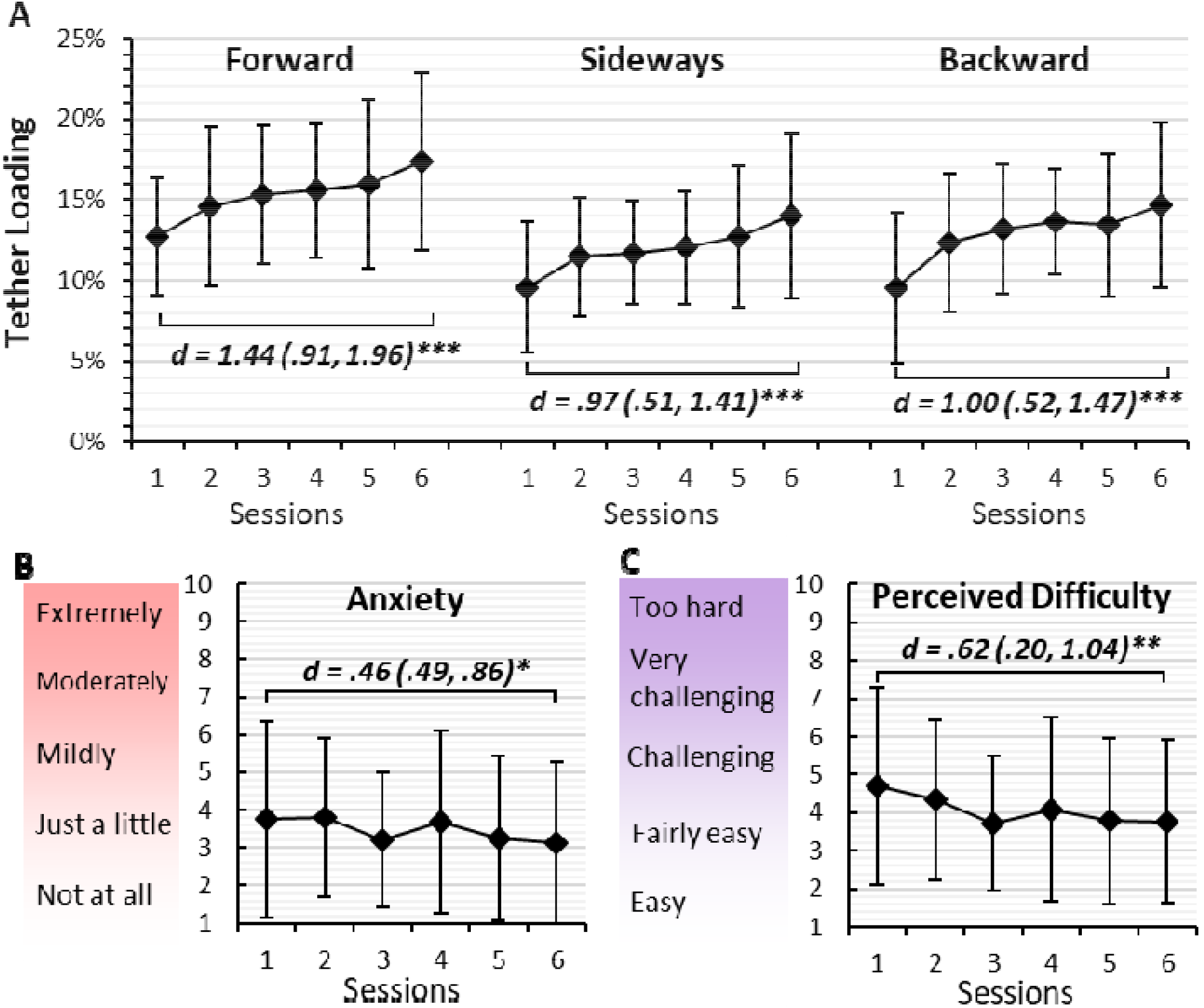
Changes in (A) maximum successful tether loading in the forward, sideways and backward reactive stepping, (B) anxiety and (C) perceived difficulty ratings during the ReacStep training. Mean +/-standard deviation. Cohen’s d (95% confidence interval) indicates small (.2), medium (.5) and large (.8) effects. *P <.05, **P <.01, *** P <.001

## DISCUSSION

This study evaluated the efficacy of the ReacStep program in improving balance recovery after trips and slips and reducing fall risk factors in older adults. Our primary hypothesis was not supported, as there was no significant difference in laboratory-induced falls during unexpected trips and slips while walking between the intervention and control groups at post-test. However, our secondary hypothesis was confirmed ReacStep improved usual gait speed, step reaction time, and maximum step length. The program also achieved excellent adherence (90%), was associated with only mild to moderate anxiety that decreased over time, and received high enjoyment ratings, indicating strong acceptability among older adults.

### Task-specific improvements, but no transfer to fall reduction

The improvements observed in tether-release reactive stepping during training align with the previous studies reporting task-specific gains following repeated perturbations (5, 23, 24) likely reflecting proactive adaptations (25). Tether-release training involves reactive stepping with progressively increasing task intensity (i.e., greater leaning force necessitating quicker and larger compensatory steps). Enhanced volitional stepping capacity was evidenced by increased maximum step length and reduced step reaction time, both important fall risk factors (26). However, the non-significant reduction in laboratory-based falls suggests limited skill transfer that can be attributable to several factors.

First, tether-release training was performed in standing and involved a single reactive step, whereas trips occur during walking and involve obstruction of the swing foot, providing a direct sensory trigger that elicits either a lowering or elevating strategy of the ipsilateral foot (27, 28). Sensory feedback during foot contact with a tripping obstacle is critical for initiating a timely reactive response (29, 30), but this was absent during lumbar tether release, which does not provide the same immediate, localised cue as that provided when striking a foot-level obstacle. Consequently, tether-release training represents a part-practice approach, focusing solely on reactive stepping without addressing two essential components: (i) the absence of the foot striking an obstacle, which serves as a sensory trigger, and (ii) reactive stepping during walking tasks (31, 32). This likely contributed to the lack of effective skill transfer to a more real-life context as assessed on the trip and slip walkway.

Similarly, volitional slip training consisted of only one or two steps, enabling participants to proactively optimise their centre of mass (CoM) position forward and stiffen the sliding limb (23, 33). These adaptations were unlikely to be present in everyday slip scenarios. There may also be important kinematic differences compared to recovery from unexpected forward slips, which typically require stepping backwards and planting a foot to stop the slip. Moreover, the neuromuscular system often adapts through trial-and-error learning across varied specific tasks, a process that ReacStep did not allow, thereby limiting its generalisation to an unexpected slip while walking (34, 35).

An unexpected finding was the higher frequency of trip encounters in daily life among the intervention group compared to controls. Two factors may explain this. (i) the ReacStep group showed reduced FaB scores, indicating less cautious behaviour and potentially greater exposure to hazards; and (ii) heightened awareness of trips and slips may have led participants to recognise and report near-falls that might otherwise go unnoticed (36). While near-falls typically predict future falls (37), no significant increase in reported falls was observed, possibly reflecting improved ability to manage trips and slips. This aligns with evidence that fall prevention training can boost confidence, sometimes leading to riskier behaviours (38). These findings highlight the need for a multi-faceted approach to fall prevention, including education on environmental risk awareness and safe mobility behaviours (39).

### Acceptability and safety

This study demonstrated excellent acceptability of the ReacStep program among older adults, with a 90% attendance rate, 92% adherence to home exercises, and a progressive reduction in anxiety across sessions. Although training intensities, such as leaning load and slip distance, increased over time, the tether-release direction and slip direction/timing remained visible and predictable. This likely enhanced participants’ confidence by maintaining a reactive component in a psychologically manageable way. These findings are consistent with our previous survey which reported high acceptability of the ReacStep program (9).

Safety outcomes were favourable. Only three participants (7%) reported pain flare-ups related to pre-existing conditions, and just one case (2%) resulted in withdrawal. Muscle strains occurred during trip/slip exposure and strength assessments, not during training sessions. These findings are encouraging given the inherently high-risk nature of RBT, particularly when compared to a previous systematic review that reported adverse events in 29% of participants in RBT groups (7).

### Study limitations

Several limitations should be acknowledged. First, although most participants expressed concern about falling, a potential healthy volunteer and high socio-economic status bias may limit the generalisability of the findings to the broader older population. Second, the study did not achieve its intended sample size, increasing the risk of a Type II (β) error. Third, although the control group received home-based exercises, participants were not blinded to group allocation, so a placebo effect cannot be ruled out. Finally, because this is a laboratory-based trial, the findings may have limited applicability to clinical and community settings.

### Future directions

While practising segmented trip and slip responses in a standing position may aid familiarisation— particularly for older adults with frailty or fear of falling—progression to walking-based perturbations is likely essential to enhance ecological validity. Effective trip recovery generally requires physical obstruction of the swing foot, activating sensory and proprioceptive systems to improve reactive responses (27, 28). For slips, meaningful neuromuscular adaptations appear to depend on stepping and postural reactions triggered by unexpected slips during walking. Future research should incorporate these task-specific components into training programs while ensuring that perturbation methods remain low-cost, acceptable, safe, and widely accessible. This approach will maximise impact and scalability, including in low-resource settings.

## Conclusion

The ReacStep program had limited impact on reactive balance during unexpected trips, slips, and real-life falls. However, it demonstrated excellent adherence and acceptability and produce improvements in gait and the ability to take larger and quicker steps, both recognised fall risk factors in older adults. Beyond its primary outcomes, this study generated valuable insights for optimising RBT, which can inform both clinical implementation and future research design. These findings have the potential to guide clinicians and researchers in developing more effective and targeted RBT interventions.

## Data Availability

Requests for data access must be directed to Dr Yoshiro Okubo and will be reviewed by the UNSW Human Research Ethics Committee (HREC).

## References

1. Gerards MHG, McCrum C, Mansfield A, Meijer K. Perturbation-based balance training for falls reduction among older adults: Current evidence and implications for clinical practice. Geriatr Gerontol Int. 2017;17(12):2294–303.

2. Okubo Y, Schoene D, Lord SR. Step training improves reaction time, gait and balance and reduces falls in older people: a systematic review and meta-analysis. Br J Sports Med. 2017;51(7):586–93.

3. Pai YC, Bhatt T, Yang F, Wang E. Perturbation training can reduce community-dwelling older adults’ annual fall risk: a randomized controlled trial. J Gerontol A Biol Sci Med Sci. 2014;69(12):1586–94.

4. McCrum C, Gerards MHG, Karamanidis K, Zijlstra W, Meijer K. A systematic review of gait perturbation paradigms for improving reactive stepping responses and falls risk among healthy older adults. Eur Rev Aging Phys Act. 2017;14:3.

5. Okubo Y, Sturnieks DL, Brodie MA, Duran L, Lord SR. Effect of Reactive Balance Training Involving Repeated Slips and Trips on Balance Recovery Among Older Adults: A Blinded Randomized Controlled Trial. J Gerontol A Biol Sci Med Sci. 2019;74(9):1489–96.

6. Mansfield A, Danells CJ, Inness EL, Musselman K, Salbach NM. A survey of Canadian healthcare professionals’ practices regarding reactive balance training. Physiother Theory Pract. 2021;37(7):787–800.

7. Devasahayam AJ, Farwell K, Lim B, Morton A, Fleming N, Jagroop D, et al. The Effect of Reactive Balance Training on Falls in Daily Life: An Updated Systematic Review and Meta-Analysis. Phys Ther. 2022;103(1).

8. Mansfield A, Aqui A, Danells CJ, Knorr S, Centen A, DePaul VG, et al. Does perturbation-based balance training prevent falls among individuals with chronic stroke? A randomised controlled trial. BMJ Open. 2018;8(8):e021510.

9. Ho C, Sharma S, Huang T, Cheung D, Hicks C, Treacy D, et al. Clinician acceptability of the ReacStep reactive balance training program for fall prevention. Physiotherapy Research International. 2024;29(4).

10. Allin LJ, Brolinson PG, Beach BM, Kim S, Nussbaum MA, Roberto KA, et al. Perturbation-based balance training targeting both slip- and trip-induced falls among older adults: a randomized controlled trial. BMC Geriatrics. 2020;20(1).

11. Pijnappels M, Reeves ND, Maganaris CN, van Dieen JH. Tripping without falling; lower limb strength, a limitation for balance recovery and a target for training in the elderly. J Electromyogr Kinesiol. 2008;18(2):188–96.

12. Rogers MW, Creath RA, Gray V, Abarro J, McCombe Waller S, Beamer BA, et al. Comparison of Lateral Perturbation-Induced Step Training and Hip Muscle Strengthening Exercise on Balance and Falls in Community-Dwelling Older Adults: A Blinded Randomized Controlled Trial. J Gerontol A Biol Sci Med Sci. 2021;76(9):e194–e202.

13. Yang F, Pai YC. Automatic recognition of falls in gait-slip training: Harness load cell based criteria. J Biomech. 2011;44(12):2243–9.

14. Duncan RP, McNeely ME, Earhart GM. Maximum Step Length Test Performance in People With Parkinson Disease: A Cross-sectional Study. J Neurol Phys Ther. 2017;41(4):215–21.

15. Lord JP, Aitkens SG, McCrory MA, Bernauer EM. Isometric and isokinetic measurement of hamstring and quadriceps strength. Arch Phys Med Rehabil. 1992;73(4):324–30.

16. Lord SR, Sambrook PN, Gilbert C, Kelly PJ, Nguyen T, Webster IW, et al. Postural stability, falls and fractures in the elderly: results from the Dubbo Osteoporosis Epidemiology Study. Med J Aust. 1994;160(11):684-5, 8-91.

17. Llinas-Regla J, Vilalta-Franch J, Lopez-Pousa S, Calvo-Perxas L, Torrents Rodas D, Garre-Olmo J. The Trail Making Test. Assessment. 2017;24(2):183–96.

18. Benedict RH, DeLuca J, Phillips G, LaRocca N, Hudson LD, Rudick R, et al. Validity of the Symbol Digit Modalities Test as a cognition performance outcome measure for multiple sclerosis. Mult Scler. 2017;23(5):721–33.

19. Benedict RHB, Schretlen D, Groninger L, Brandt J. Hopkins Verbal Learning Test – Revised: Normative Data and Analysis of Inter-Form and Test-Retest Reliability. The Clinical Neuropsychologist. 1998;12(1):43–55.

20. Clemson L, Bundy AC, Cumming RG, Kay L, Luckett T. Validating the Falls Behavioural (FaB) scale for older people: a Rasch analysis. Disabil Rehabil. 2008;30(7):498–06.

21. Lamb SE, Jorstad-Stein EC, Hauer K, Becker C, Prevention of Falls Network E, Outcomes Consensus G. Development of a common outcome data set for fall injury prevention trials: the Prevention of Falls Network Europe consensus. J Am Geriatr Soc. 2005;53(9):1618–22.

22. Pavol MJ, Owings TM, Foley KT, Grabiner MD. Mechanisms leading to a fall from an induced trip in healthy older adults. J Gerontol A Biol Sci Med Sci. 2001;56(7):M428–37.

23. Bhatt T, Yang F, Pai Y-C. Learning to Resist Gait-Slip Falls: Long-Term Retention in Community-Dwelling Older Adults. Archives of Physical Medicine and Rehabilitation. 2012;93(4):557–64.

24. Dijkstra BW, Horak FB, Kamsma YP, Peterson DS. Older adults can improve compensatory stepping with repeated postural perturbations. Front Aging Neurosci. 2015;7:201.

25. Bohm S, Mademli L, Mersmann F, Arampatzis A. Predictive and Reactive Locomotor Adaptability in Healthy Elderly: A Systematic Review and Meta-Analysis. Sports Med. 2015;45(12):1759–77.

26. Okubo Y, Schoene D, Caetano MJ, Pliner EM, Osuka Y, Toson B, et al. Stepping impairment and falls in older adults: A systematic review and meta-analysis of volitional and reactive step tests. Ageing Res Rev. 2021;66:101238.

27. Phu S, Sturnieks DL, Song PYH, Lord SR, Okubo Y. Treadmill induced belt-accelerations may not accurately evoke the muscle responses to obstacle trips in older people. J Electromyogr Kinesiol. 2024;75:102857.

28. Eng JJ, Winter DA, Patla AE. Strategies for recovery from a trip in early and late swing during human walking. Exp Brain Res. 1994;102(2):339–49.

29. Benjuya N, Melzer I, Kaplanski J. Aging-induced shifts from a reliance on sensory input to muscle cocontraction during balanced standing. J Gerontol A Biol Sci Med Sci. 2004;59(2):166–71.

30. Bloem BR, Allum JH, Carpenter MG, Honegger F. Is lower leg proprioception essential for triggering human automatic postural responses? Exp Brain Res. 2000;130(3):375–91.

31. Rhein Z, Vakil E. Motor sequence learning and the effect of context on transfer from part-to-whole and from whole-to-part. Psychol Res. 2018;82(3):448–58.

32. Wang Y, Wang S, Bolton R, Kaur T, Bhatt T. Effects of task-specific obstacle-induced trip-perturbation training: proactive and reactive adaptation to reduce fall-risk in community-dwelling older adults. Aging Clin Exp Res. 2020;32(5):893–905.

33. Pavol MJ, Runtz EF, Pai YC. Young and older adults exhibit proactive and reactive adaptations to repeated slip exposure. J Gerontol A Biol Sci Med Sci. 2004;59(5):494–502.

34. Mount J, Pierce SR, Parker J, DiEgidio R, Woessner R, Spiegel L. Trial and error versus errorless learning of functional skills in patients with acute stroke. NeuroRehabilitation. 2007;22(2):123–32.

35. Nunes J, Armada M, Pereira JL, Ribeiro NF, Carvalho O, Santos CP. Biomechanical strategies for mitigating unexpected slips: A review. J Biomech. 2024;173:112235.

36. Siegmund GP, Heiden TL, Sanderson DJ, Inglis JT, Brault JR. The effect of subject awareness and prior slip experience on tribometer-based predictions of slip probability. Gait Posture. 2006;24(1):110–9.

37. Tiedemann A, Purcell K, Clemson L, Lord S, Sherrington C. Fall prevention behaviour after participation in the Stepping On program: a pre–post study. Public Health Research & Practice. 2021;31(1).

38. Quach L, Galica AM, Jones RN, Procter-Gray E, Manor B, Hannan MT, et al. The nonlinear relationship between gait speed and falls: the Maintenance of Balance, Independent Living, Intellect, and Zest in the Elderly of Boston Study. J Am Geriatr Soc. 2011;59(6):1069–73.

39. Cheung D, Paul SS, Mackenzie L, Wesson J, Goh L, Canning CG, et al. A scoping review of safe mobility behaviour in fall prevention: implications for people with Parkinson’s disease. Disabil Rehabil. 2025;47(13):3278–91.

