## Supplementary file for "Effects of the ‘Reac Step’ training program on balance recovery and fall risk factors in older people: An assessor-blinded randomised controlled trial"

| Supplementary Table 1 - Comparison between participants who completed and dropped out from the study | | | |
| --- | --- | --- | --- |
| Variables | Completed (N=75) | Dropped (N=13) | P |
| Age (years) | 72.1 (5.5) | 74.0 (8.1) | 0.448 |
| Gender (female) | 73% (55) | 54% (7) | 0.155 |
| Height (cm) | 165.9 (8.9) | 170.4 (9.3) | 0.096 |
| Weight (kg) | 71.0 (13.4) | 77.6 (18.5) | 0.128 |
| Body mass index (kg/m^2^) | 25.8 (4.4) | 26.7 (6.0) | 0.591 |
| Osteoporosis (yes) | 18% (13) | 0% (0) | 0.342 |
| Back pain (yes) | 21% (15) | 11% (1) | 0.679 |
| Hip pain (yes) | 17% (12) | 22% (2) | 0.653 |
| Knee pain (yes) | 31% (22) | 0% (0) | 0.057 |
| Feet pain (yes) | 17% (12) | 0% (0) | 0.342 |
| Anxiety disorder (yes) | 7% (4) | 0% (0) | 1.000 |
| Use of walking stick (yes) | 3% (2) | 0% (0) | 1.000 |
| Fear of falling (yes) | 81% (61) | 75% (9) | 0.696 |
| Falls in the past year (1+ falls) | 45% (34) | 33% (4) | 0.436 |

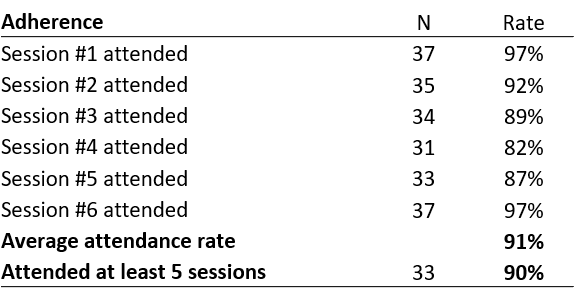

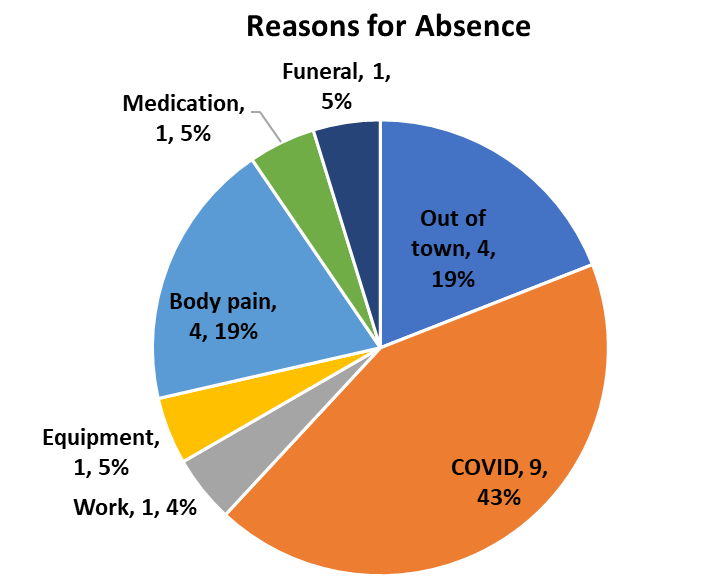

Supplementary Figure 1 - Reasons for absence from the ReacStep training sessions. A total of 21 absence by 14 participants.

Five participants who withdrew from the study prior to session #1 are not included.

Supplementary Table 2 - Adherence to ReacStep training sessions (N=38).

| Supplementary Table 3 - Adherence to home-based strength training (N=75) | | | | |
| --- | --- | --- | --- | --- |
| Variables | Intervention group  (N=37) | | Control group  (N=38) | *P* |
| Week 1 | 1.8 | ± 0.5 | 1.9 ± 0.8 | 0.207 |
| Week 2 | 2.1 | ± 0.9 | 2.1 ± 0.5 | 0.987 |
| Week 3 | 2.0 | ± 0.9 | 2.0 ± 0.5 | 0.997 |
| Week 4 | 2.2 | ± 1.1 | 2.2 ± 0.5 | 0.870 |
| Week 5 | 2.0 | ± 0.5 | 1.9 ± 0.6 | 0.561 |
| Week 6 | 2.0 | ± 0.4 | 2.0 ± 0.7 | 0.846 |
| Week 7 | 2.0 | ± 0.4 | 1.9 ± 0.7 | 0.850 |
| Week 8 | 1.7 | ± 0.8 | 1.9 ± 0.8 | 0.422 |
| Average | 2.0 | ± 0.4 | 2.0 ± 0.4 | 0.765 |
| Values are means ± standard deviations of strength training sessions per week. Each session included banded squat, hip flexors (right/left), hip abductors (right/left) and hip extensors (right/left) for 10 repetitions each using an appropriate resistance band. Participants were instructed to conduct 2 sessions each week during the study period. | | | | |

**Feedback related to the ReacStep program from the intervention participants**

- “I'm glad to be involved in this research and trust that the benefits are far-reaching. May I suggest an advertising campaign to alert the public of the importance of maintaining strength training, in particular balance exercises. Hopefully this would target the ageing population. Thank you all for your dedicated work.”
- “The fit of the harness often varied in the weekly training sessions i.e., it felt either more or less secure - perhaps a variable that didn't influence outcome.”
- “Thank you, I became less nervous about falling, good training.”
- “Undergoing the study has made me more aware of my surroundings and how I feel about ageing. I now feel so much more confident about going about my daily activities as I was beginning to lose that (confidence).”
- “I really enjoyed the whole program. At first, I was a bit nervous about the training. But the ReacStep Study Team were very patient with me, and I finally lost my nervousness and began to enjoy myself. Thank you to all of you in the team.”
- “All the training involved stepping over an object or slipping on an object, yet when the final test occurred, the wooden trapdoor was so high and opened in such a way that it was impossible to step over and instead I tripped so badly that I almost went flying!”
- “I would be interested in access to some kind of maintenance program to ensure that motor skills learned do not dissipate but get entranced in reflexes.”
